# The smallest worthwhile effect on pain and function for rotator cuff repair surgery: a benefit-harm trade-off study

**DOI:** 10.1101/2024.07.24.24310953

**Authors:** Harrison J Hansford, Rachelle Buchbinder, Joshua R Zadro, James H McAuley, Manuela L Ferreira, Adriane Lewin, Richard S Page, Ian A Harris

## Abstract

**Background:** The smallest worthwhile effect (SWE) is the minimum benefit required in addition to that from a comparator, for an intervention to be considered worthwhile by patients. We aimed to estimate the SWE for rotator cuff repair (with decompression and debridement) compared to either decompression and debridement alone or to non-surgical treatment for people with atraumatic shoulder pain.

**Methods:** Benefit-harm trade-off study. We recruited English speaking adults aged 45-75 years with shoulder pain of intensity ≥4 (on a 0-10 scale) for ≥6 months to our online survey through paid advertising on Facebook. Participants must have sought care in the past 6-months and could not have had recent shoulder surgery or significant recent shoulder trauma. Participants were explained three treatments: rotator cuff repair (with subacromial decompression and debridement), subacromial decompression and debridement alone and non-surgical treatment. Participants completed the benefit-harm trade-off survey to determine the SWE of improvements in pain and function for rotator cuff repair compared to the other treatments and again after one week to assess reliability. We used univariable linear regression to estimate associations between baseline characteristics and SWE.

**Results:** We recruited 56 participants. The mean ± standard deviation age was 58.4±6.7 years and 39 (70%) were female. For rotator cuff repair to be worthwhile compared to decompression and debridement alone participants needed to see at least a median 40% (interquartile range (IQR) 20-62.5) between-group improvement in pain and function.

Compared to non-surgical treatment, the SWE was a median 40% (IQR 30-60). On the Western Ontario Rotator Cuff (WORC) Index the SWE values equate to a between-group improvement of 28/100 points (533/2100 on the raw WORC score). Female sex was associated with larger SWEs for both comparisons. Reliability analyses were underpowered, 25/56(45%) provided follow-up data; the intraclass correlation coefficient estimates ranged from 0.60-0.77.

**Conclusions:** This SWE indicates the benefit required by people with shoulder pain to consider the costs and risks of surgical rotator cuff repair worthwhile is larger than previously estimated minimum clinically important differences (13.5-28/100 on the WORC Index). This SWE may be used to inform the design or interpret the findings of trials of these comparisons.

## Introduction

The lifetime prevalence of shoulder pain is estimated to be up to 67% and this prevalence increases with age(1). Rotator cuff disease is commonly identified as the cause of shoulder pain(2). For people with shoulder pain attributable to rotator cuff disease with or without a rotator cuff tear, most clinical practice guidelines recommend initial non-surgical treatment such as simple analgesia, exercise therapy and glucocorticoid injection (3). For people with persisting symptoms that fail to improve over time, surgery is often recommended (4).

Recommended surgeries may include subacromial decompression and debridement with or without repair of the rotator cuff for those with tears (5, 6).

There is moderate certainty evidence that subacromial decompression and debridement alone provides minimal benefit over placebo or ongoing non-surgical care (6). Consequently, this procedure is no longer recommended by some guidelines (7) and is declining in use (8). There is less certainty about the value of rotator cuff repair as there are no high-quality trials comparing this to placebo repair and it does not appear beneficial over non-operative care (6). To determine the true value of rotator cuff repair we are currently performing such a trial (the Australian Rotator Cuff Trial - ACTRN12620000789965). However, to inform design and interpretation of this trial, as well as clinical care, we need to understand the extent to which patients consider the potential risks and inconveniences of rotator cuff repair to be worthwhile compared to subacromial decompression and debridement alone (i.e. placebo control), or non-surgical treatment. This is currently unknown (6).

The smallest worthwhile effect is a threshold used to determine clinical importance from a patient’s perspective. (9-12). The smallest worthwhile effect is the minimum benefit that would have to be achieved with an intervention to make its costs, risks, and inconveniences worthwhile, compared to some alternative (e.g. no treatment, other interventions) (9, 13). It is often determined using the benefit-harm trade-off method, which involves outlining to participants the relevant attributes of an intervention including the costs, risks, and inconveniences, before asking a series of questions about the minimum benefit they would require to make the intervention worthwhile (9-11). Despite being commonly used for this purpose (11, 14-17), the reliability of the estimates derived from benefit-harm trade-off has never been investigated.

The primary aim of this study was to determine the smallest worthwhile effect for two comparisons of treatments for atraumatic shoulder pain attributable to rotator cuff disease. The first compared rotator cuff repair (including decompression and debridement) with decompression and debridement alone, and the second compared rotator cuff repair (including decompression and debridement) with non-surgical treatment. Our secondary aim was to determine the test-retest reliability of the benefit-harm trade-off method for determining the smallest worthwhile effect. We aim to use this finding to inform the sample size calculation of the Australian Rotator Cuff Trial.

## Materials and methods

This was a benefit-harm trade-off study. The study protocol (available via the Open Science Framework (OSF) at osf.io/crj9p) was completed prior to data collection. We report findings consistent with STROBE (18). We received ethical approval from the University of New South Wales Human Research Ethics Committee (HC230374). The survey codebook and all deidentified data and code used for the analysis is available on OSF.

### Inclusion criteria

Participants were eligible if they were 45-75 years of age inclusive, proficient in the English language, and reported having persisting shoulder pain. For the purposes of this study, persisting shoulder pain was defined as having shoulder pain of intensity ≥ four out of ten on a numerical pain rating scale (NPRS, where 0 is ‘no pain’ and 10 is ‘worst possible pain’) (19), for at least the past six months. We only included participants if they had seen a health professional for their shoulder pain and had received some form of active non-surgical treatment such as medication, exercise therapy and/or glucocorticoid injection within this time period. We excluded participants if they had recent surgery (less than six months ago) to the affected shoulder or were living outside of Australia, due to different costs and treatment protocols globally. We also excluded participants who had injured their shoulder in a traumatic event (i.e., more than a simple fall from standing height). We excluded participants if they reported being told they have a rotator cuff tear > 4cm, or inflammatory or systemic arthritis. The eligibility criteria ensured that participants were similar to those eligible for the currently recruiting Australian Rotator Cuff trial (ACTRN12620000789965) and similar to patients who are offered rotator cuff repair in routine clinical practice.

### Recruitment

Participants were recruited to our survey by paid advertisements on social media (Facebook) and by trial surgeons. When participants opened the online survey, they had an opportunity to read the participant information sheet which detailed the research project. If participants chose to complete the survey, informed consent was implied.

### Outcomes and data collection

Using Research Electronic Data Capture (REDCap) (20, 21) we collected demographic and descriptive data including: sex, age, diagnosis for shoulder pain (i.e., rotator cuff tear, ‘bursitis’, etc.), clinician who made the diagnosis, previous shoulder surgery, history of active non-surgical treatment (i.e., reported having tried physical therapy including exercise therapy, medication, and glucocorticoid injection), pain intensity over the past 24 hours using the 0-10 NPRS (19), duration of shoulder pain, type of occupation and physical activity levels. We also included the health subscale of a risk-taking questionnaire (22) and whether they would consider shoulder surgery for their pain, answered with a ‘yes’ or ‘no’. The complete survey is available at (osf.io/crj9p).

### Benefit-harm trade-off

Participants were presented with a written description of each intervention explaining the costs, potential risks, and inconveniences (Supplementary Text 1). For the comparator intervention (i.e., non-surgical treatment or shoulder decompression and debridement) it was outlined that each of these treatments would, on average, result in a 20% improvement in symptoms within 6 months. This was the change associated with each treatment as reported in a recent review (6), and clinical trial (23) respectively. Participants were shown two benefit-harm trade-off comparisons (Supplementary Text 1). Prior to this study, these descriptions were pilot-tested with two consumers with lived experience of shoulder pain via individual 30-minute video-calls and their feedback was incorporated to improve understanding of what was being asked.

After reading the outlines of the intervention (rotator cuff repair), and comparator (either non-surgical treatment or shoulder decompression and debridement alone), participants were asked whether they would consider rotator cuff repair worthwhile if they were to achieve total recovery. If they said they would not consider rotator cuff repair worthwhile, they were not required to complete the benefit-harm trade-off survey as they were not considered to be representative of the target population, i.e., patients considering having a surgical rotator cuff repair.

Participants who indicated they would consider the rotator cuff repair worthwhile were then asked to rate the percentage improvement in their pain and function they would require, in addition to the 20% improvement expected from either comparator, to make the rotator cuff repair worthwhile (11, 14). This outcome was chosen as pain and function are important outcomes for patients and are used to determine the need for surgery (24). Following this, they were asked whether they would still consider the intervention worthwhile if the improvement in symptoms was 10% less than the value they had indicated. This process was continued in increments of 10% until the participant no longer considered rotator cuff repair worthwhile. The final improvement in pain and function to which participants responded ‘yes’ was deemed the smallest worthwhile effect for that participant. The order in which the comparisons were displayed to participants was pseudo-randomised based on record ID to limit any potential effects of the ordering of the questions.

To assist with interpretation of our results we converted the percentage of the smallest worthwhile effect to a between-group point-estimate on the Western Ontario Rotator Cuff (WORC) Index, based on the baseline WORC values in this sample. The WORC Index is a 21-item measure of shoulder pain and function with scores from 0-2100 (25). While higher scores on the index represent greater impairment, the scale is often transformed to a 0-100 scale where lower scores represent greater impairment. We calculated point estimates on both the untransformed and transformed scale. The WORC index is the primary outcome of the Australian Rotator Cuff trial. Point-estimates of the smallest worthwhile effect were calculated as in Equation 1 using several hypothetical baseline WORC index values, including the baseline of the present sample.

Equation 1: Calculation of point-estimates of the smallest worthwhile effect.

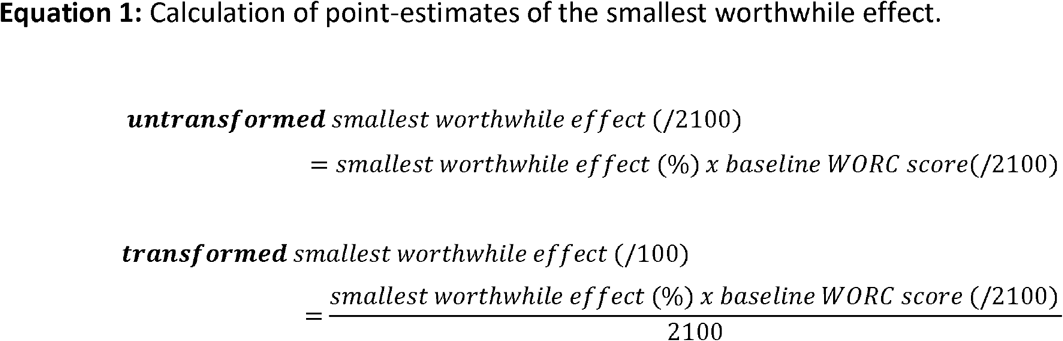

### Reliability

We also sought to determine the test-retest reliability of the benefit-harm trade-off method. To do this, we sent an automated email to study participants one week after completing the initial benefit-harm trade-off questions, asking them to re-take the same questions.

### Sample size calculation

There are no sample size guidelines for estimating the smallest worthwhile effect. For the assessment of univariable associations between individual baseline variables and the smallest worthwhile effect with a significance level of alpha = 0.05 and power 80% required a sample of n = 56. The sample required to estimate the reliability of the benefit-harm trade-off method using a test-retest design was n = 53 (26). This sample size was calculated using a minimum acceptable reliability of 0.7, an expected reliability of 0.85, with a significance level of alpha = 0.05, power 80%.

### Data analysis

We conducted all analyses using R Statistical Software (27). We summarised descriptive data using mean and standard deviation, or median and interquartile range for data that were not normally distributed. Data normality was inspected visually. The overall smallest worthwhile effect for each comparison was reported as the median value of each individual smallest worthwhile effect. We present the interquartile range of the smallest worthwhile effect. We used univariable linear regression models to estimate associations between baseline characteristics (sex, pain intensity, age, baseline WORC index, duration of symptoms, perceived health status, physical demands of occupation, health related risk taking, and level of exercise) and the smallest worthwhile effect for each comparison. We used the non-parametric bootstrap with 1000 replicates to estimate 95% confidence intervals (CI) for the observed associations (28). Baseline associations are presented on the scale of the smallest worthwhile effect (i.e., % improvement in pain and function). To determine the reliability of the benefit-harm trade-off method we calculated the interclass correlation coefficient (ICC) two-way random effects, absolute agreement, single rater/measurement (ICC(2,1)) (29, 30) of the two ratings of the smallest worthwhile effect.

## Results

642 people accessed the initial survey link, 585 consented to complete the survey, 83 of whom were eligible. Of those eligible, 56 participants provided data for at least one analysis (67% completion rate). Characteristics of participants included in the study are presented in Table 1.

**Table 1.**
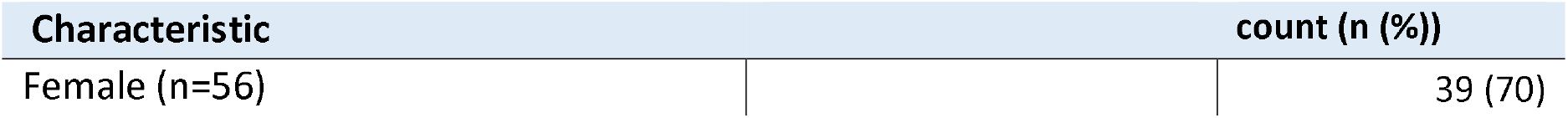
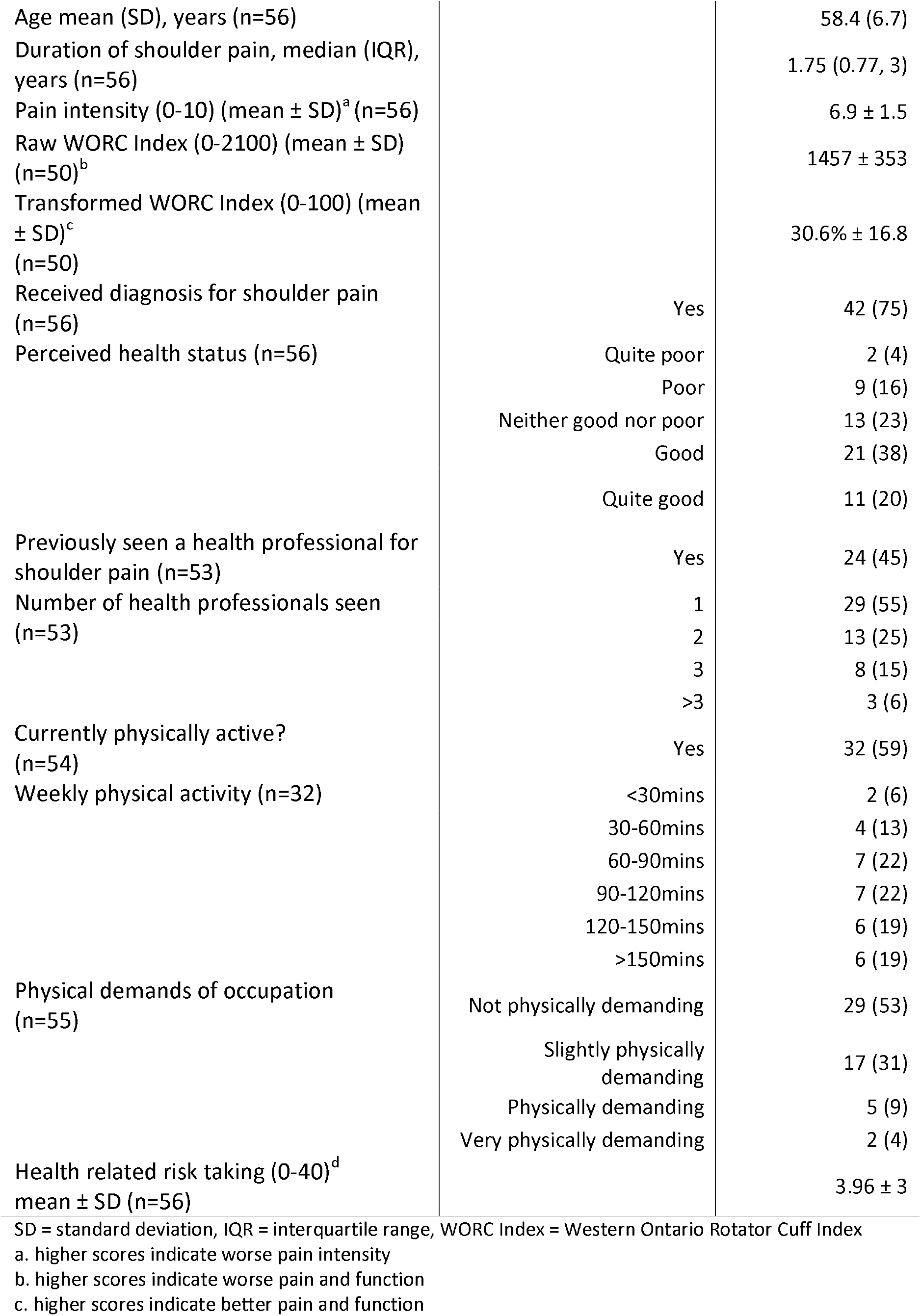
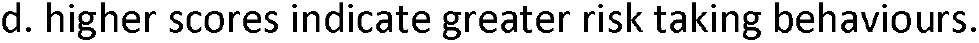
Characteristics of participants who contributed to at least one analysis (n=56)

| Characteristic | count (n (%)) |
| --- | --- |
| Female (n=56) | 39 (70) |

|  |  |  |
| --- | --- | --- |
| Age mean (SD), years (n=56) |  | 58.4 (6.7) |
| Duration of shoulder pain, median (IQR), years (n=56) |  | 1.75 (0.77, 3) |
| Pain intensity (0-10) (mean ± SD)<sup>a</sup> (n=56) | | 6.9 ± 1.5 |
| Raw WORC Index (0-2100) (mean ± SD) (n=50)<sup>b</sup> | | 1457 ± 353 |
| Transformed WORC Index (0-100) (mean ± SD)<sup>c</sup> (n=50) | | 30.6% ± 16.8 |
| Received diagnosis for shoulder pain (n=56) | Yes | 42 (75) |
| Perceived health status (n=56) | Quite poor | 2 (4) |
|  | Poor | 9 (16) |
|  | Neither good nor poor | 13 (23) |
|  | Good | 21 (38) |
|  | Quite good | 11 (20) |
| Previously seen a health professional for shoulder pain (n=53) | Yes | 24 (45) |
| Number of health professionals seen (n=53) | 1 | 29 (55) |
|  | 2 | 13 (25) |
|  | 3 | 8 (15) |
|  | >3 | 3 (6) |
| Currently physically active? (n=54) | Yes | 32 (59) |
| Weekly physical activity (n=32) | <30mins | 2 (6) |
|  | 30-60mins | 4 (13) |
|  | 60-90mins | 7 (22) |
|  | 90-120mins | 7 (22) |
|  | 120-150mins | 6 (19) |
|  | >150mins | 6 (19) |
| Physical demands of occupation (n=55) | Not physically demanding | 29 (53) |
|  | Slightly physically demanding | 17 (31) |
|  | Physically demanding | 5 (9) |
|  | Very physically demanding | 2 (4) |
| Health related risk taking (0-40)<sup>d</sup> mean ± SD (n=56) | | 3.96 ± 3 |

SD = standard deviation, IQR = interquartile range, WORC Index = Western Ontario Rotator Cuff Index
a. higher scores indicate worse pain intensity
b. higher scores indicate worse pain and function
c. higher scores indicate better pain and function
d. higher scores indicate greater risk taking behaviours.

### The smallest worthwhile effect of rotator cuff repair compared to decompression and debridement alone

17/53 (31%) participants reported they would not consider rotator cuff repair for their shoulder pain. For participants with shoulder pain who would consider rotator cuff repair (n=36), the smallest worthwhile effect was a 40% (IQR 20% to 60%) improvement in pain and function, additional to decompression and debridement (Figure 3). There were several baseline characteristics that were associated with a greater smallest worthwhile effect. Females required a 22.3%, 95% CI 8.7 to 36.1 greater smallest worthwhile effect, as did those who had a greater self-perception of health (7.1%, 95% CI 0.1 to 14.9) and were more physically active (3.8%, 95% CI 0.2 to 7.4). No other meaningful associations were observed between the baseline characteristics and the smallest worthwhile effect (Supplementary Table 1).

**Figure 3.**
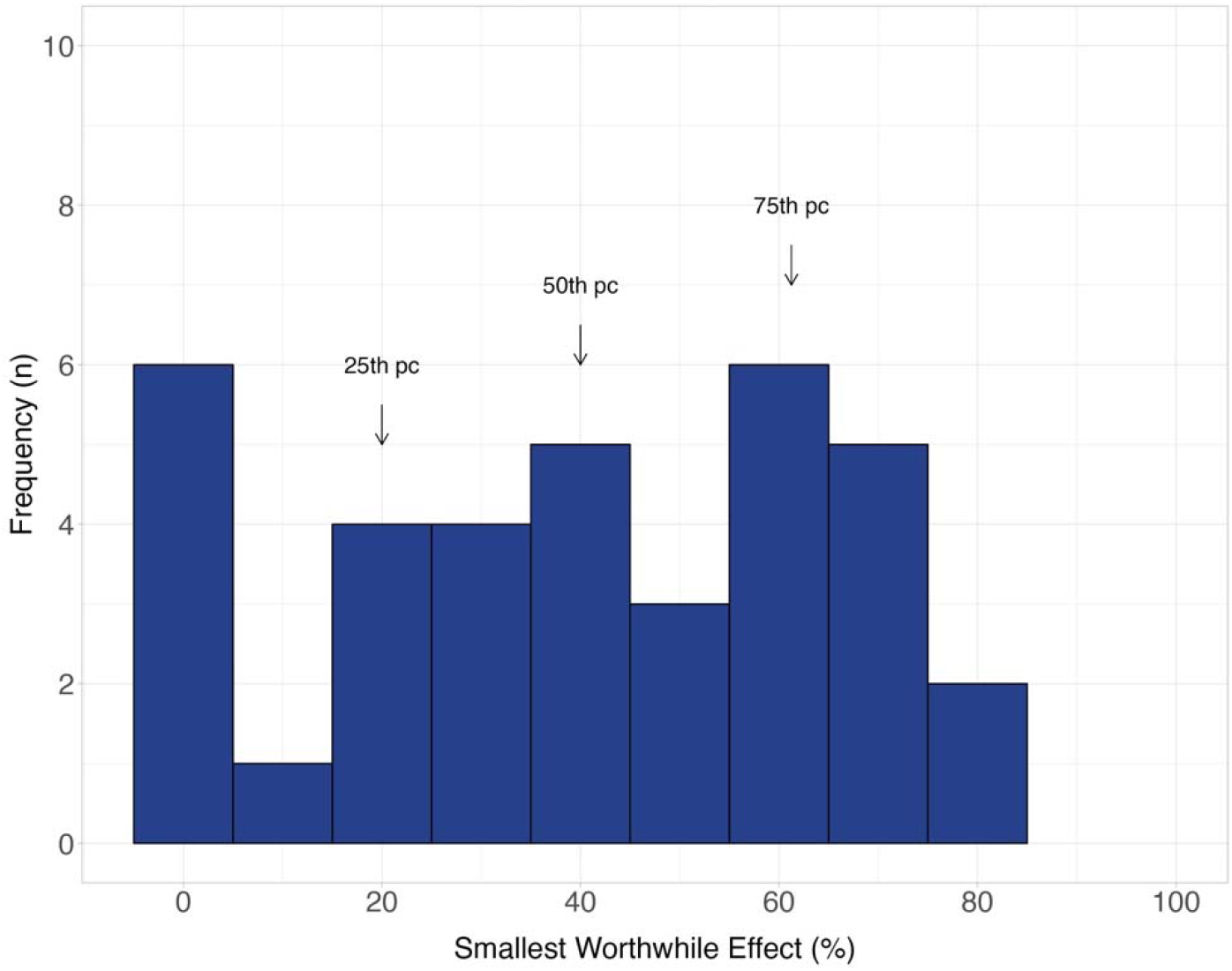
Distribution of the smallest worthwhile effect of pain and function for rotator cuff repair compared to decompression and debridement (n=36)

**Figure 4.**
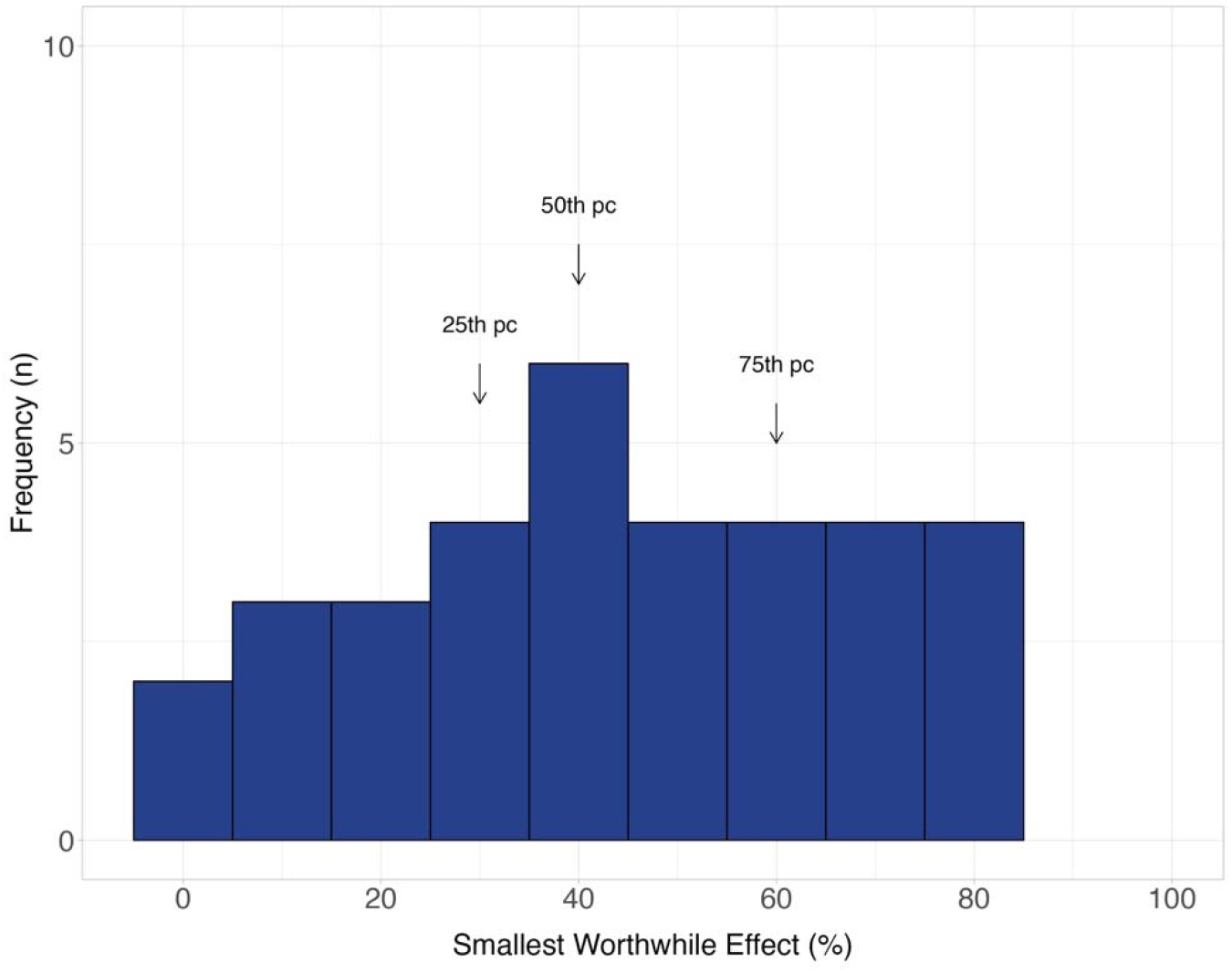
Distribution of the smallest worthwhile effect of pain and function for rotator cuff repair compared to non-surgical treatment (n=34)

### The smallest worthwhile effect of rotator cuff repair compared to non-surgical treatment

17/51 (33%) participants reported they would not consider rotator cuff repair for their shoulder pain. For people who would consider rotator cuff repair (n=34), the smallest worthwhile effect was a 40% (IQR 30% to 60%) improvement in pain and function, additional to non-surgical treatment (Figure 3). Participants who were female required a 14.6% (95% CI 0.8 to 27.5) larger smallest worthwhile effect. No other meaningful associations were observed between the baseline characteristics and the smallest worthwhile effect (Supplementary Table 2).

Table 2 outlines how the relative smallest worthwhile effect corresponds with point-estimates on the WORC scale at different hypothetical levels of baseline pain and function. See Supplementary Text 2 for further worked examples of the calculation of the point-estimates.

**Table 2:** Point estimates of the smallest worthwhile effect corresponding with different hypothetical levels of baseline pain and function.

| Hypothetical baseline pain and function | Between-group difference required for rotator cuff repair to be considered worthwhile compared to decompression & debridement or non-surgical treatment |
| --- | --- |

| WORC raw (0-2100)<sup>a</sup> | WORC Transformed (0-100)<sup>b</sup> | Untransformed (0-2100) | Transformed (0-100) |
| --- | --- | --- | --- |
| 1680 | 20 | 672 | 32 |
| 1457† | 30.6 | 588 | 28 |
| 1260 | 40 | 504 | 24 |
| 1050 | 50 | 420 | 20 |

WORC = Western Ontario Rotator Cuff Index, SWE = smallest worthwhile effect
† score in the present sample
a. higher scores indicate worse pain and function
b. higher scores indicate better pain and function

### Reliability

25/56 (45%) completed at least one follow-up question. This was significantly less than the required sample to assess the test-retest reliability (n=53). For the reliability of the benefit-harm trade-off comparison of rotator cuff surgery compared to decompression and debridement alone, 24 participants completed follow-up. In this sample, 7 (29%) participants did not consider rotator cuff surgery worthwhile, even with a complete improvement in pain and function. In the follow-up benefit-harm trade-off, 8 (33%) participants did not consider rotator cuff surgery worthwhile. Of the 16 participants who would consider surgery in the follow-up, the intra-class correlation coefficient (ICC) two-way random effects, absolute agreement, single rater/measurement (ICC(2,1)) was moderate to good, ICC = 0.76 (95% CI = 0.53 to 0.89) (31).

For the reliability of the benefit-harm trade-off comparison of rotator cuff surgery compared to non-surgical treatment, 25 participants completed follow-up. In this sample, 7 (28%) participants did not consider rotator cuff surgery worthwhile, even with a complete improvement in pain and function. In the follow-up benefit-harm trade-off, 5 (25%) participants did not consider rotator cuff surgery worthwhile. Of the 20 participants who would consider surgery in the follow-up, the ICC(2,1) was poor to good, ICC = 0.60 (95% CI = 0.23 to 0.82) (31).

## Discussion

In our benefit-harm trade-off study with 56 participants, the smallest worthwhile effect was an additional 40% improvement in pain and function compared with either subacromial decompression and debridement alone, or non-surgical treatment. Based on the baseline WORC index score of our sample, the 40% smallest worthwhile effect equates to a between-group difference of 588 on the raw WORC score, or 28 points when transformed to the 0-100 scale of the WORC Index. Notably, being female was associated with greater improvement required to consider rotator cuff repair worthwhile compared to males, for both comparators (decompression and debridement alone and non-surgical treatment).

The minimum clinically important difference (MCID) is another threshold of clinical importance. The MCID for the WORC has been estimated between 282.6 to 588.7/2100 (13.5 to 28/100) (32), lower values than our estimates of SWE. Our estimates of the smallest worthwhile effect provide estimates of the minimum benefit that patients would consider rotator cuff repair worthwhile compared to either debridement and decompression alone or non-surgical treatment. These estimates of the smallest worthwhile effect can be used to interpret the results of relevant clinical trials assessing the efficacy of rotator cuff repair compared to decompression and debridement alone - a comparison currently being evaluated by our team (ACTRN12620000789965); and non-surgical interventions previously investigated (6).

The smallest worthwhile effect has several notable benefits compared to the MCID: it is determined directly by patients, it represents a between-group difference rather than a within-group change and is treatment-comparison specific (9, 12). The MCID is usually estimated irrespective of treatment comparison and applied as a property of an instrument, rather than of the comparison (33). Surgery is costly and can involve significant risks and inconveniences for patients. Therefore, it could be expected that the treatment effect required by patients to consider surgery worthwhile would be larger than other, less invasive interventions and therefore is likely to be larger than the MCID. Our study demonstrates this indirectly as the smallest worthwhile effect of 40% is qualitatively larger than other estimates of the smallest worthwhile effect for non-surgical interventions for musculoskeletal pain such as exercise (20%) and nonsteroidal anti-inflammatory drugs (30%) in low back pain (11, 15, 34). Our estimates of the smallest worthwhile effect are similar to those for total knee replacement(35), strengthening the notion that patients require greater improvements from more invasive treatments for them to be considered worthwhile. Using the MCID to estimate the importance of surgical treatment effects may underestimate the improvement patients require to consider them worthwhile.

This smallest worthwhile effect study is being used to power a randomised trial of shoulder surgery comparing rotator cuff repair with debridement and decompression to debridement and decompression alone (ACTRN12620000789965). To our knowledge, the smallest worthwhile effect has not been used in this way previously. One approach to using the smallest worthwhile effect to power a study is to calculate the point-estimate that equates to the percentage improvement in symptoms (e.g., 40% with a transformed baseline WORC of 30/100 (a raw score of 1470/2100) is a between-group difference of 28/100 on the WORC (a raw difference of 588/2100)). This point estimate can then be used in a regular power calculation. This approach may lead to smaller trials that do not subject additional participants to potentially unnecessary risks. This approach is comparable statistically to using a smaller (MCID-based) value because if a trial is powered to detect a larger effect but is unable to do so, it is unlikely patients will consider the intervention worthwhile, even if the effect was more precise.

### Strengths and limitations

A relatively small number of participants completed the benefit-harm trade-off questions when compared to previous studies conducted by our group(11, 34). This was due to the larger proportion of participants who would not consider rotator cuff repair for their pain and function, even if it offered complete improvement. This may be because our sample was not presenting to surgery, or that surgery is a treatment that has greater costs, risks, and inconveniences compared to others that have been compared for painful conditions, such as physiotherapy care, exercise and nonsteroidal anti-inflammatory drugs (11, 15, 34). Due to a low follow-up rate (25/56, 45%) we suggest interpreting our results of the reliability analyses with caution.

Further, as our study mimicked the eligibility criteria of the Australian Rotator Cuff trial, there were a significant number of consenting people who were not eligible to participate. While this provides an informative smallest worthwhile effect for the Australian Rotator Cuff trial, it may limit the generalisability of these findings to all patients with shoulder pain due to other causes. However, a key difference between our inclusion criteria and the Australian Rotator Cuff trial is that we did not assess whether participants had a rotator cuff tear, however, we assumed they may consider treatments similarly to those who did. By transforming the smallest worthwhile effect to the WORC scale, we are implying that the weighting of pain and function that participants gave when answering the benefit-harm trade-off question will approximate the weighting of the WORC. We chose to assess the smallest worthwhile effect as a global score of pain and function, rather than the WORC to limit the cognitive burden of the survey.

To our knowledge, this is the first benefit-harm trade-off study to compare active treatments, rather than an active treatment to no treatment. This may increase the relevance of the estimated smallest worthwhile effects to policy makers and study interpretations as it is less typical to compare treatments to no treatment in the field of pain due to the impact of contextual effects of interventions(36).

## Conclusions

The smallest worthwhile effect for rotator cuff repair was an additional 40% improvement in pain and function when separately compared to subacromial decompression and debridement, and non-surgical interventions. In our sample, this smallest worthwhile effect equates to a between-group difference of 28/100 points on the WORC Index (a raw difference of 588/2100). This smallest worthwhile effect should be used to interpret clinical trials and systematic reviews of shoulder rotator cuff repair compared to decompression and debridement, or non-surgical interventions.

## Data Availability

The survey codebook is available on OSF (osf.io/crj9p) All de-identified data and code used for the analysis will be made available upon publication on OSF

