## Supplementary Material for "The smallest worthwhile effect on pain and function for rotator cuff repair surgery: a benefit-harm trade-off study"

**Appendices**

**Supplementary Text 1**: Example descriptions of interventions.

**Supplementary Table 1:** Associations between baseline variables and smallest worthwhile effect for rotator cuff surgery compared to decompression and debridement alone

**Supplementary Table 2:** Associations between baseline variables and smallest worthwhile effect for rotator cuff surgery compared to non-surgical treatment

**Supplementary Text 2**: worked examples of translating % smallest worthwhile effect to Western Ontario Rotator Cuff Index Scores

**Appendix 1**: Example descriptions of interventions.

Comparison 1: Shoulder surgery with rotator cuff repair compared to shoulder surgery without rotator cuff repair

We are trying to find out how effective shoulder rotator cuff repair surgery would have to be at improving your shoulder symptoms to make it worthwhile compared to another shoulder surgery that is similar but does not touch the rotator cuff. Imagine you saw your doctor and they mentioned that two of the following treatments were available to improve your shoulder symptoms.

The first procedure, rotator cuff repair is a surgical procedure that involves the surgeon making small skin cuts in your shoulder (keyhole surgery), removing part of the bone and soft tissue to broaden the tendon passage (decompression) and **repair of the torn tendons**. After the surgery you will be out of hospital in the same day, you'll have your arm in a sling for three to six weeks and will not be able to drive for that time. You will be recommended to attend physiotherapy sessions to gradually improve your function over 6 months.

In the other surgery without the rotator cuff repair, you will undergo a similar procedure, however, you will only have the surgery to remove part of the bone and soft tissue to broaden the passage of the tendon (decompression), reducing the time taken to perform the surgery, the time it takes to recover from surgery, and reducing the chance of complications from surgery. You will also be out of hospital the same day. You will have your arm in a sling for a shorter time, between 2 days and 2 weeks, you will be able to return to work and drive within 2 weeks and you may have 2-3 months of physiotherapy to improve your function.

The risks of the surgery with rotator cuff repair that are greater than the other surgery include prolonged stiffness and tendon re-tear post-surgery.

Below is a summary of the above information

|  | Rotator cuff surgery | Decompression surgery |
| --- | --- | --- |
| Inconveniences | · Shoulder in sling for 3 to 6 weeks  · Unable to drive for 3 to 6 weeks  · May take up to 12 weeks for you to perform daily activities or work that involves lifting above your head or heavy objects.  · Physical rehabilitation for 3 to 12 months post-surgery involving attendance at outpatient clinic every 2 to 4 weeks. | · Shoulder in sling for 2 days to 2 weeks  · Able to work and drive within 2 weeks  · Physical rehabilitation for 1 to 3 months post-surgery involving attendance at outpatient clinic every 2 to 4 weeks. |
| Costs | · Free through public hospital - 1 year wait  · $0-$13,000 through private hospital depending on your level of private health insurance | · Similar costs, although if not covered by private health insurance it may be $1000 cheaper  · i.e., $0-$12,000 |
| Risks | · Risk prolonged stiffness and re-tearing the tendon post-surgery  · Small surgical risks of blood clots, infection, or excessive bleeding. | · Only small surgical risks of blood clots, infection, or excessive bleeding. |

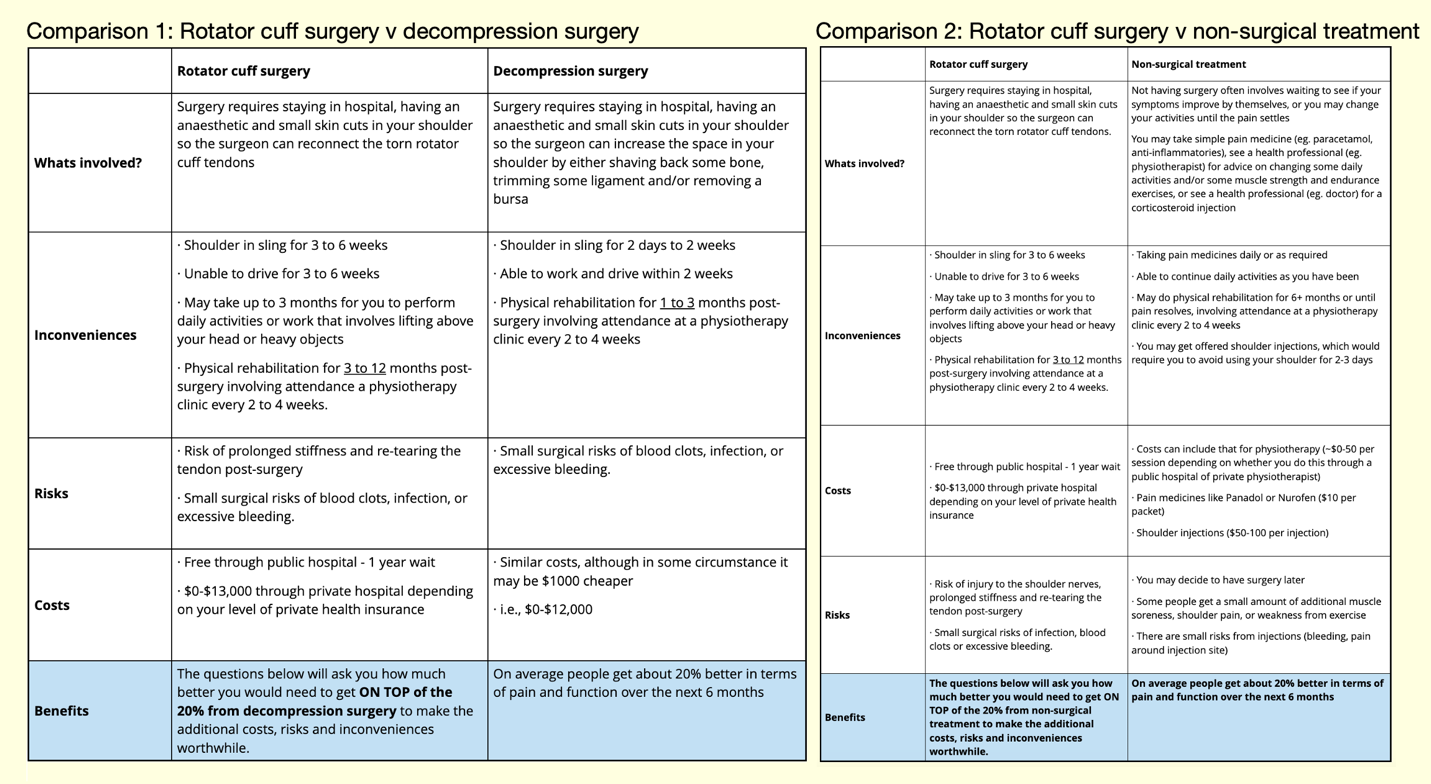

Tabulated Benefit-harm trade-off comparisons as displayed to survey participants after each detailed description.

Comparison 2: Rotator cuff surgery compared to non-surgical treatment

We are trying to find out how effective shoulder rotator cuff repair surgery would have to be at improving your shoulder symptoms to make it worthwhile compared to non-surgical treatment, which mostly includes exercise, pain medicines, and sometimes shoulder injections. Imagine you saw your doctor and they mentioned that two of the following treatments were available to improve your shoulder symptoms.

Rotator cuff repair is a surgical procedure that involves the surgeon making small incisions in your shoulder (keyhole surgery), removing part of the bone and soft tissue to broaden the tendon passage (subacromial decompression) and repair of the torn tendons. After the surgery you will be out of hospital the same day, you'll have your arm in a sling for three to six weeks and will not be able to drive for that time. You will be recommended to attend physiotherapy sessions at an outpatient clinic every 2 to 4 weeks to gradually improve your function over 6 months.

There are some risks of the surgery including infection, excessive bleeding a small risk of blood clots. People who have this procedure may also experience injury to the shoulder nerves, prolonged stiffness and re-tearing the tendon post-surgery. If you have the surgery through a public hospital, there are no out of pocket expenses, although the wait time can be up to a year. If you go through a private hospital, depending on your private health cover, you may be out of pocket as little as $0 if you have private health insurance with full hospital cover and no excess, or up to $12,000 if you have no private health insurance.

If you do not choose to have surgery, the most likely course of action would be to maintain non-surgical treatment, which might include some strengthening exercise for your shoulder, simple analgesic medicines as required (i.e., Panadol or Nurofen) and sometimes you may be recommended to have a shoulder injection to reduce the pain.

If you do not have the surgery, your ability to perform your day to day activities could improve by about 20% over the next 6 months.

Below is a summary of the above information

|  | Rotator cuff surgery | Non-surgical treatment |
| --- | --- | --- |
| Inconveniences | · Shoulder in sling for 3 to 6 weeks  · Unable to drive for 3-6 weeks  ·  May take up to 12 weeks for you to perform daily activities or work that involves lifting above your head or heavy objects.  · Physical rehabilitation for 3-12 months post-surgery involving attendance at outpatient clinic every 2 to 4 weeks. | · Time to attend health appointments (1hr + travel time every fortnight) |
| Costs | · Free through public hospital - 1 year wait  · $0-$13,000 through private hospital depending on your level of private health insurance | · Costs can include that for physiotherapy (~$0-50 per session depending on whether you do this through a public hospital of private physiotherapist)  · Pain medicines like Panadol or Nurofen ($10 per packet)  · Shoulder injections ($50-100 per injection) |
| Risks | · Risk of injury to the shoulder nerves, prolonged stiffness and re-tearing the tendon post-surgery  · Small surgical risks of infection, blood clots or excessive bleeding. | · You may decide to have surgery later  · Some people get a small amount of muscle soreness, shoulder pain, or weakness |

**Appendix 2:** Associations between baseline variables and smallest worthwhile effect for rotator cuff surgery compared to decompression and debridement alone

Supplemental Table 1. Smallest worthwhile effect of rotator cuff surgery compared to decompression and debridement alone (n=36)

| Baseline characteristic | Association (ß [95% CI]) |
| --- | --- |
| Sex (Female = 1) | 22.3% [8.7 to 36.1]* |
| Pain intensity (0-10) | -2.2% [-9.5 to 4.9] |
| Age (years) | -0.3% [-1.6 to 1.0] |
| WORC Index (0-100) | 0.2% [-0.2 to 0.9] |
| Duration of symptoms (years) | 0.4% [-1.0 to 2.3] |
| Perceived health status (0-5) | 7.1% [0.1 to 14.9]* |
| Physical demands of occupation (0-4) | -3.5% [-16.0 to 8.5] |
| Health related risk taking (0-32) | -1.0% [-2.9 to 1.8] |
| Level of exercise | 3.8% [0.22 to 7.35]* |

Note: ß is on the scale of the smallest worthwhile effect, i.e., percent improvement in pain and function

**Appendix 3:** Associations between baseline variables and smallest worthwhile effect for rotator cuff surgery compared to non-surgical treatment

Supplemental Table 2. Smallest worthwhile effect of rotator cuff surgery compared to decompression and debridement alone (n=34)

| Baseline characteristic | Association (ß [95% CI]) |
| --- | --- |
| Sex (Female = 1) | 14.6% [0.8 to 27.5]* |
| Pain intensity (0-10) | -1.8% [-5.1 to 8.6] |
| Age (years) | 0.6% [-0.6 to 1.7] |
| WORC Index (0-100) | 0.2% [-0.3 to 0.8] |
| Duration of symptoms (years) | 0.4% [-0.3 to 2.7] |
| Perceived health status (0-5) | 2.9% [-4.5 to 9.8] |
| Physical demands of occupation (0-4) | -5.5% [-16.8 to 5.0] |
| Health related risk taking (0-32) | -1.0% [-2.7 to 1.9] |
| Level of exercise | 1.9% [-9.6 to 9.0] |

Note: ß is on the scale of the smallest worthwhile effect, i.e., percent improvement in pain and function

**Appendix 4**: worked examples of translating % smallest worthwhile effect to Western Ontario Rotator Cuff Index Scores

Patient cohort starts at a score of 1440, which is equivalent to 30% function

Briefly this can be calculated as:

1440*0.4 = 576

= 576/2100

= 27.4% between groups

Illustrated fully

First calculate 60% (i.e., smallest worthwhile effect + 20% change due to comparator) of raw score / 2100

e.g., 60% x 1440 = 864

which brings the endpoint to = 1440 – 864 = 576 / 2100

which equates to a % of (2100 – 576)/2100 = 72.5%

Calculate the change associated with comparator (20% of raw score)

e.g., 20% x 1440 = 288

which brings the endpoint to = 1440 – 288 = 1152 / 2100

which equates to a % of (2100 – 1152)/2100 = 45.1%

The smallest worthwhile effect between groups is therefore 576 points, or 27.4% on the percentage scale

**If the baseline score of the cohort is higher (i.e., the people have greater function)**

Cohort starts at a score of 1050, which is 50% function

Briefly this can be calculated as:

1050*0.4 = 420

= 420/2100

= 20% between groups

First calculate 60% of raw score / 2100

e.g., 60% x 1050 = 630

which brings the endpoint to = 1050 – 630 = 420 / 2100

which equates to a % of (2100 – 420)/2100 = 80%

Calculate the change associated with comparator (20% of raw score)

e.g., 20% x 1050 = 210

which brings the endpoint to = 1050 – 210 = 840 / 2100

which equates to a % of (2100 – 840)/2100 = 60%

The smallest worthwhile effect between groups is therefore 420 points, or 20% on the percentage scale
